# Reliability and Performance of the IRRAflow® System for Intracranial Lavage and Evacuation of Hematomas - A Technical Note

**DOI:** 10.1101/2023.07.07.23292372

**Authors:** Mette Haldrup, Mojtaba Nazari, Chenghao Gu, Mads Rasmussen, Stig Dyrskog, Claus Ziegler Simonsen, Mads Grønhøj, Frantz Rom Poulsen, Naveed Ur Rehman, Anders Rosendal Korshoej

**Affiliations:** Department of Neurosurgery, Aarhus University Hospital, Palle Juul-Jensens Boulevard 165, 8200 Aarhus N; Department of Engineering, Electrical and Computer Engineering, Aarhus University, Finlandsgade 22, 8200, Aarhus N; Department of Anesthesiology, Gødstrup Regional Hospital, Herning, Denmark; Department of Clinical Medicine, Aarhus University, Palle Juul-Jensens Boulevard 99, 8200 Aarhus N; Department of Anesthesiology, Section of Neuroanesthesia, Aarhus University Hospital, Aarhus, Denmark; Department of Intensive Care, Aarhus University Hospital, Palle Juul-Jensens Boulevard 165, 8200 Aarhus N; Department of Neurology, Aarhus University Hospital, Palle Juul-Jensens Boulevard 165, 8200 Aarhus N; Department of Neurosurgery, Odense University Hospital, J.B. Winsløws Vej 4, 5000 Odense, Clinical Institute and BRIDGE (Brain Research - Inter Disciplinary Guided Excellence), University of Southern Denmark, Odense, Denmark

## Abstract

**Background:** Intraventricular hemorrhage (IVH) is a severe condition with poor outcomes and high mortality. IRRA*flow®* (IRRAS AB) is a new technology introduced to accelerate IVH clearance by minimally invasive wash-out. The IRRA*flow®* system performs active and controlled intracranial irrigation and aspiration with physiological saline, while simultaneously monitoring and maintaining a stable intracranial pressure (ICP). We addressed important aspects of the device implementation and intracranial lavage.

**Method:** To allow versatile investigation of multiple device parameters, we designed an *ex vivo* lab setup. We evaluated 1) compatibility between the IRRA*flow®* catheter and the Silverline f10 bolt (Spiegelberg), 2) the physiological and hydrodynamic effects of varying the IRRA*flow®* settings, 3) the accuracy of the IRRA*flow®* injection volumes, and 4) the reliability of the internal ICP monitor of the IRRA*flow®*.

**Results:** The IRRA*flow®* catheter was not compatible with Silverline bolt fixation, which was associated with leakage and obstruction. Design space exploration of IRRA*flow®* settings revealed that a balanced saline influx and efflux required adjustments of the drainage bag to adapt to different irrigation rates. High irrigation rates could be compensated by lowered drain bag height and vice versa. Appropriate settings included irrigation rate 20 ml/h with a drainage bag height at 0 cm, irrigation rate 90 ml/h with a drainage bag height at 19 cm and irrigation rate 180 ml/h with a drainage bag height at 29 cm. We found the injection volume performed by the IRRA*flow®* to be stable and reliable, while the internal ICP monitor was compromised in several ways due to the proximal location of the pressure sensor within the cassette of the device rather than within the parenchyma of the brain. Furthermore, we observed a significant mean drift difference of 3.16 mmHg (variance 0.4, p=0.05) over a 24-hour test period with a mean 24-hour drift of 3.66 mmHg (variance 0.28) in the pressures measured by the IRRA*flow®* compared to 0.5 mmHg (variance 1.12) in the Raumedic measured pressures.

**Conclusion:** Bolting of the IRRA*flow®* catheter using the Medtronic Silverline® bolt is not recommendable. Increased irrigation rates are recommendable followed by a decrease in drainage bag level. ICP measurement using the IRRA*flow®* device was unreliable and should be accompanied by a control ICP monitor device in clinical settings.

## Introduction

Intraventricular hemorrhage (IVH) is a severe condition with poor outcomes and high mortality rates(1, 2). Currently, IVH treatment relies on passive drainage of cerebral spinal fluid (CSF) with external ventricular drainage (EVD) to decrease the intracranial pressure (ICP) and facilitate hematoma evacuation^3,4^. Recent studies have shown that clot removal could be accelerated using intraventricular fibrinolysis (IVF) leading to improved survival and functional outcomes(1, 2). However, no significant treatment advances have been made in the past decades, and minimally invasive techniques to achieve better management are warranted.

A new technology, IRRA*flow®* (IRRAS AB), was introduced to accelerate IVH evacuation by minimally invasive wash-out. The IRRA*flow®* system performs active and controlled intracranial irrigation and aspiration with physiological saline while monitoring and maintaining a stable ICP. The IRRA*flow®* system is FDA-approved (2019) and CE-marked (2018) for the treatment of IVH and has currently been used to treat more than 600 patients worldwide (post-market surveillance, courtesy of IRRAS AB). Currently, there are only two published case reports evaluating IRRA*flow®* in the treatment of IVH (DRIFT and Hummingflow(3, 4). Four randomized controlled trials and one case-control study have presently been initiated to test IRRA*flow®* treatment of IVH in different settings (registered trials: ARCH (NCT05118997), ACTIVE (NCT05204849), Unregistered trials: VASH, AFFECT, and DIVE).

The IRRA*flow®* control unit has several adjustable settings, including irrigation rate, drain valve opening pressure, bolus volume, and drainage bag level. However, there are no current recommendations for best-practice treatment settings, and no studies have investigated the physiological effects of varying these parameters. IRRA*flow®* uses an internal ICP sensor connected to the catheter irrigation line to exert supervisory control over the system’s operational features, including irrigation and drainage. Documentation of sensor reliability is therefore crucial for safe device operation. To our knowledge, no former studies have evaluated these aspects of the IRRA*flow®* system to ensure patient safety and validate grounds for clinical decision-making using the technology.

In this paper, we addressed these gaps in the current knowledge on IRRA*flow®* implementation and IVH lavage using an *ex vivo* model and data from patients treated in a randomized, controlled safety/feasibility study of IRRA*flow®* performance (Active Study (NCT05204849). Specifically, we 1) conducted an extensive exploration of the physiological impact of different settings of the IRRA*flow®* control unit (design space exploration); 2) evaluated and described the data output from IRRA*flow®; 3)* conducted extensive validation and reliability tests of IRRA*flow®* volume injections and ICP readouts. Our experiments shed important light on the physiological effects and feasibility of different treatment implementations and propose a basis for clinical guidelines in the treatment of IVH using IRRA*flow®*.

## Materials and Methods

### The IRRAflow*®* device

IRRA*flow®* is an advanced drainage system that combines gravity-driven drainage with periodic irrigation and ICP monitoring. The system operates in cycles beginning with a 1-second irrigation phase, followed by a 9-second ICP measurement phase, and ends with an aspiration phase with variable duration (10 to 170 seconds) defined by the user (Fig. 1).

**Figure 1.**
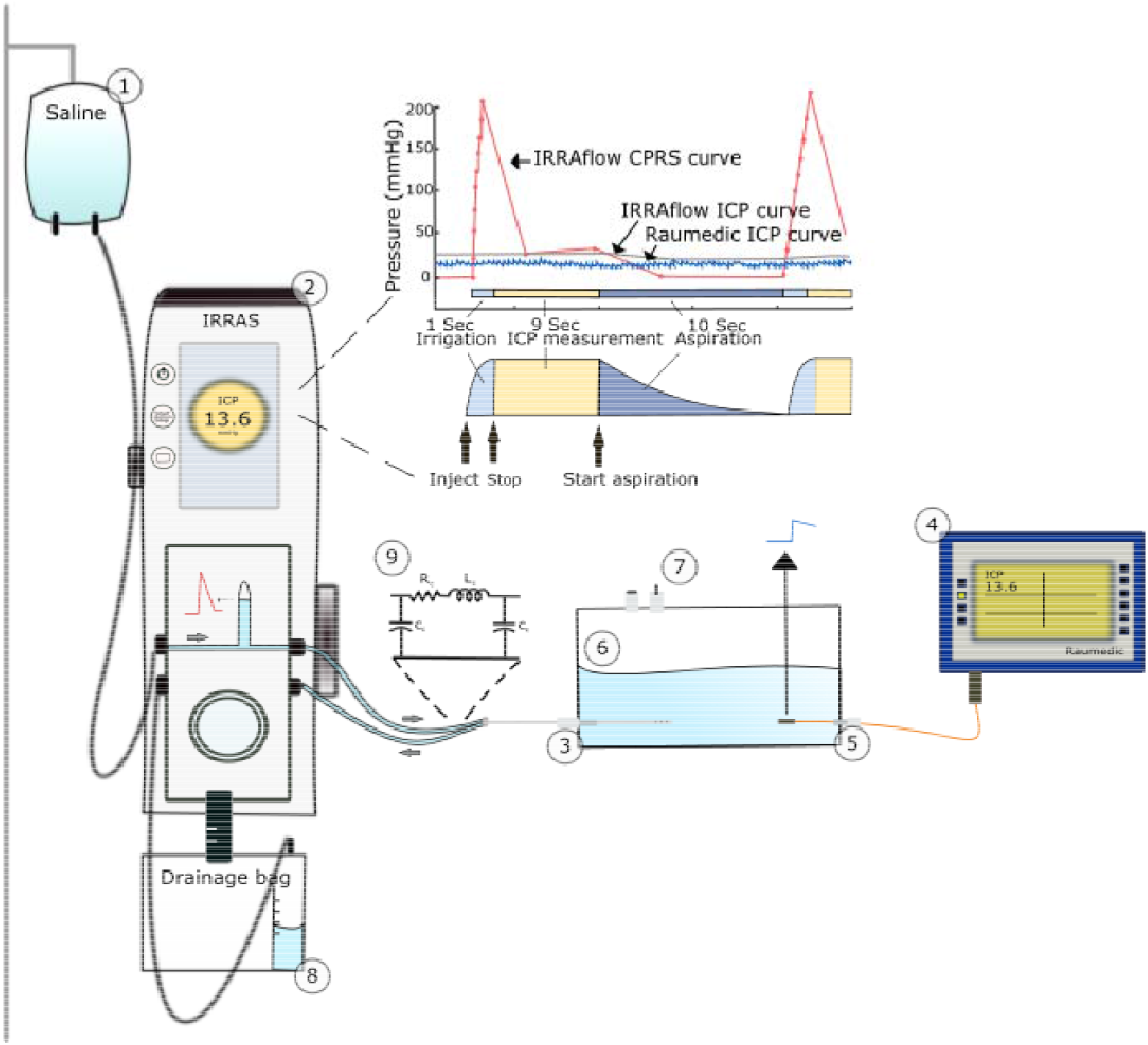
Lab setup and pressure signals. 1. Saline bag, 2. IRRA*flow®* device, 3. Bolt-connected dual lumen IRRA*flow®* catheter, 4. Raumedic device for ICP monitoring, 5. Raumedic ICP monitor probe, 6. Acrylic chamber, 7. Valves, 8. Drainage bag, 9. schematic illustration of the proximal pressure sensing in the internal IRRAflow*®* ICP monitor showing a spiked CPRS signal from IRRA*flow®* compared to a synchronized and stable ICP signal obtained using the Raumedic probe at the distal catheter tip. The IRRA*flow®* current pressure signal (CPRS, red solid line), the IRRA*flow®* ICP signal (gray solid line), and the Raumedic ICP signal (blue solid line). The CPRS shows a rapid pressure increase during the first second irrigation phase followed by a stabilizing decrease during the 9s ICP measurement phase, and finally a further decrease during the aspiration phase. The CPRS has a lower sampling rate (red dots) and a lower signal to noise ratio (SNR) compared to the Raumedic ICP signal. The IRRA*flow®* ICP curve represents a single point value in time, sampled from the final CPRS value at the end of the measurement phase, when the CPRS signal has settled. This value is then maintained at a constant until the end of the next measurement phase. The RLC circuit is shown above the irrigation line (9). (R, resistance; L, inductance; C, compliance). Furthermore, the cycle operations of the IRRAflow*®* are illustrated.

The system uses a peristaltic pump to deliver 0.5 or 1 ml of saline infusion over 1 second during the irrigation phase. Users can set the cumulative irrigation rates between 20 ml/hour and 180 ml/hour depending on the duration of drainage and hence the frequency of bolus infusions.

IRRA*flow®* monitors the ICP using pressure transducers located in the cassette of the device, coupled to the tip of the catheter via the irrigation tubing of the tube set. The sensor measures ICP as a hydrostatic pressure, so it must be placed in a liquid environment to function properly. The pressure measured at the pressure transducer is equivalent to the pressure at the catheter tip when the irrigation line is fluid-filled, unobstructed, the pressure transducer is positioned at the same height as the tip of the catheter, and the liquid is still and not in motion. The decoupling of the pressure transducer from the tip of the catheter allows for the pressure transducer signal to be re-zeroed in situ. An ICP value is output to the user at the end of the 9s Measurement Phase, and this displayed average pressure is displayed throughout the Drain and Irrigation Phase until updated after the next Measurement Phase (fig. 1). The intrinsic ICP readout during the Monitoring Phase serves as supervisory control of drainage and irrigation. The measured pressure is then compared to the Drain Above Setting. If the measured pressure is lower than the Drain Above Setting, the IRRAflow System remains in Measurement Mode until the measured pressure exceeds the Drain Above Setting. If the measured pressure is higher than the Drain Above Setting, the control unit enters Drainage Mode. The IRRAflow System monitors the ICP signal throughout the entire IRRAflow cycle and is identified as the current pressure signal (CPRS), which can be displayed upon user prompt.

The Drainage Phase can vary between 10 and 170 seconds, depending on the user-set irrigation bolus volume (0.5 or 1 ml of saline infusion over approximately 1 second) and the user-set irrigation flow rate, which range from 20 ml/hour to 180 ml/hour. IRRA*flow* controls drainage via a solenoid pinch-type valve within the cassette of the device. Drainage is controlled by the user via the “Drain above” setting. When the measured pressure during the Measurement Phase exceeds the Drain Above Setting, the system enters the Drainage Phase, the solenoid pinch valve opens and liquid is allowed to drain. If the measured pressure is below the Drain Above Setting, the system enters and remains in the Measurement Phase (no drainage and no irrigation occur) until the measured pressure is greater than the Drain Above Setting.

Like a standard EVD, IRRA*flow®* drainage is gravity-driven. However, with a standard EVD, the user adjusts the level of the drainage line relative to the catheter tip to define the pressure gradient driving the aspiration. With the IRRA*flow®* the drainage is driven by the gradient between the ICP and the level of the drainage bag, when the ICP is within the operational range, which can be set from -15 cmH2O to -103 cmH2O below the catheter tip. This results in suction of CSF during drainage, with gradients ranging from 0 to -75 mmHg.

### The lab setup

To allow versatile investigation of multiple device parameters, we designed an *ex vivo* lab setup replicating the hydrodynamic conditions of the intracranial space. We used a stiff 10x10x7 cm acrylic box with a 10 mm wall thickness. We filled it with 200 ml of room-temperature physiological saline (viscosity: 0.7 mPa) and 500 ml of atmospheric air at 23 degrees Celsius to attain physiological system compliance (ΔV/ΔP) of 0.60-0.70 ml/mmHg(5). The temperature of the fluid was controlled by the Raumedic PtO2 monitor and a thermostat in the lab. Two airtight manual valves with 10 mm circular openings provided interior chamber access. To establish access points for ventricular drains, pressure sensors, and syringes, we fixed two catheter bolts (Silverline®, Spiegelberg GmbH & Co), a Raumedic Neurovent-P bolt, and a female luer-lock at 1 cm height from the chamber bottom.

We inserted a dual-lumen IRRA*flow®* catheter (version 2.0 ICGS020) 50 mm deep through the Silverline bolt and the catheter tip was placed precisely in the hight of the “0” reference on the IRRA*flow* device. The IRRA*flow®* cassette was connected to the IRRA*flow®* control unit (IRRAS, version 3.0). Tubing and saline bag attachment were prepared following the product guidelines. We inserted a Raumedic Neurovent-P pressure sensor at the height of the IRRA*flow®* catheter tip to continuously monitor the chamber pressure equivalent to the ICP (Fig. 1). The Neurovent-P sensor and the IRRA*flow®* device were calibrated and zeroed before all experiments. We measured the saline input and output on a scale before each experiment to accurately monitor the amount of irrigated saline. We set the IRRA*flow®* alarm limits to the maximum range and the Drain Above setting to -99 mmHg to maintain device operation under extreme conditions (Supplementary Material 2).

### Patient data

The *in vivo* data set for this study was collected prospectively from patients who were a part of the Active study, a randomized controlled phase 2 study testing the safety and feasibility of IRRA*flow®*. Patient recruitment occurred from January 2022 to November 2022. The data was obtained from patients with severe spontaneous primary or secondary IVH treated with IRRA*flow®*. Patients were followed for 3 month and the authors had full written permission to access patients information. All patients had a Raumedic Neurovent-P monitor placed in the same hemisphere as the IRRA*flow®* catheter to obtain correlated datasets. ICP signals were extracted from the IRRA*flow®* control unit and the Raumedic MPR-2 datalogger upon the end of IRRA*flow®* treatment and analyzed using MATLAB, MathWorks. Ethical approval was obtained from the Central Denmark Committee on Health Research Ethics (journal number 1-10-72-34-21) and participant consent was collected from legal authorized representatives for all patients in accordance with relevant Danish legislation and the Helsinki Declaration.

### Study design and hypotheses

#### Experiment 1 - Bolt compression cap tightness test

In this experiment, we aimed to evaluate the compatibility between the IRRA*flow®* catheter and the Silverline f10 bolt from Spiegelberg GmbH & Co as the IRRA*flow* is placed using a tunneling technique (Supplementary material 1). We tested the following hypotheses:

1. Leakage would occur when the bolt was only loosely tightened around the IRRA*flow®* catheter.
2. The IRRA*flow®* catheter would be increasingly constricted with an increasing number of bolt revolutions.

This was important to assess the clinical feasibility of implementing bolt mounting of IRRAflow*® and* to determine the appropriate settings of the *ex vivo* setup. To test these hypotheses, we performed five IRRA*flow®* irrigation tests of 1-hour duration for at different bolt revolutions with and without the catheter insertion stylet *in situ*. We evaluated the IRRA*flow®* irrigation performance and the integrity of the catheter fixation to determine the optimal settings, ideally preventing obstruction and leakage.

#### Experiment 2 - IRRAflow*®* Design Space Exploration

In this experiment, we aimed to evaluate the physiological and hydrodynamic effects of varying the IRRA*flow®* settings. We conducted an extensive design space exploration with comprehensive variations of the following device parameters in different combinations:

1. Irrigation rates: 20 ml/h, 90 ml/h, and 180 ml/h.
2. Drainage bag levels: +39 cm, 0 cm, -19 cm, -29 cm, -39 cm and -49 cm.

For each test, we performed five repetitions of 10-minute duration. In addition, the Drain Above parameter was set to -99 and alarm limits to -99 to 99 to ensure continuous device operation throughout the experiments.

#### Experiment 3 - Accuracy of the IRRAflow*®* Injection Volume

This experiment aimed to assess the accuracy and reliability of saline volumes injected by the IRRA*flow®* system. We hypothesized that increasing pressure within the chamber would compromise the internal saline pump, resulting in a reduced injection volume. To test this, we conducted repeated consecutive 1 ml bolus infusion cycles in a closed system, gradually increasing chamber pressure. Our observations utilized a scale measuring the injected volume in grams. IRRA*flow®* infusions were stopped when chamber pressure exceeded the upper critical limit of 99 mmHg. We repeated the experiment with different baseline chamber volumes (100-500 ml) to ensure stable results and repeated the tests 5 times at each baseline volume.

#### Experiment 4 - Accuracy and reliability of the IRRAflow*®* pressure sensor

In this experiment, we aimed to evaluate the accuracy and reliability of the internal IRRA*flow®* ICP monitor by evaluating the current pressure signal (CPRS) output of IRRA*flow®* in two different ways:

1. Assessment of the CPRS during the irrigation phase of the IRRA*flow®* operation cycle compared to the CPRS during manual injections using a syringe. A Raumedic device was used as a control unit.
2. Assessment of the CPRS during the aspiration cycle and comparison of the pressure curves with the Raumedic-derived pressure signals.

Furthermore, we measured deviations between the IRRA*flow®* ICP signals compared to the Raumedic ICP signals in both the *ex vivo* setting and in real patient data. The real patient data were extracted in a time-synchronized manner for two patients and in an unsynchronized manner for two additional patients. The distributions of the unsynchronized data were evaluated, and deviations were quantified for each patient using a two-sample t-distribution test. The null hypothesis was that there was no difference in the pressure values between the two devices.

To evaluate sensor stability, we conducted five drift tests of 24-hour duration for both IRRA*flow®* and the Raumedic ICP sensors. These tests were performed with different baseline chamber pressures (6, 8, and 10 mmHg). The drift data was analyzed using a Gardner-Altman plot and further a histogram made from which mean drift, variance, and a corresponding correlating p-value between the data from the two devices was calculated.

## Results

### IRRA*flow*® data output

The IRRA*flow®* data output was extracted using the data export function. The data structure comprised pressure curves and a variety of values (i.e., CPRS, System and treatment status, ICP, upper and lower alarm limits, drain above setting, infusion volume (ml/hour), cycle time (sec), etc.) (Supplementary material 3). The pressure readouts consisted of two variables: 1) the ICP signal, which is a constant value sampled at the end of the ICP measurement phase and maintained throughout the remaining cycle, and 2) the current pressure signal (CPRS), which gives the real-time ICP signal throughout the entire cycle (Figs. 1 and 2).

**Figure 2.**
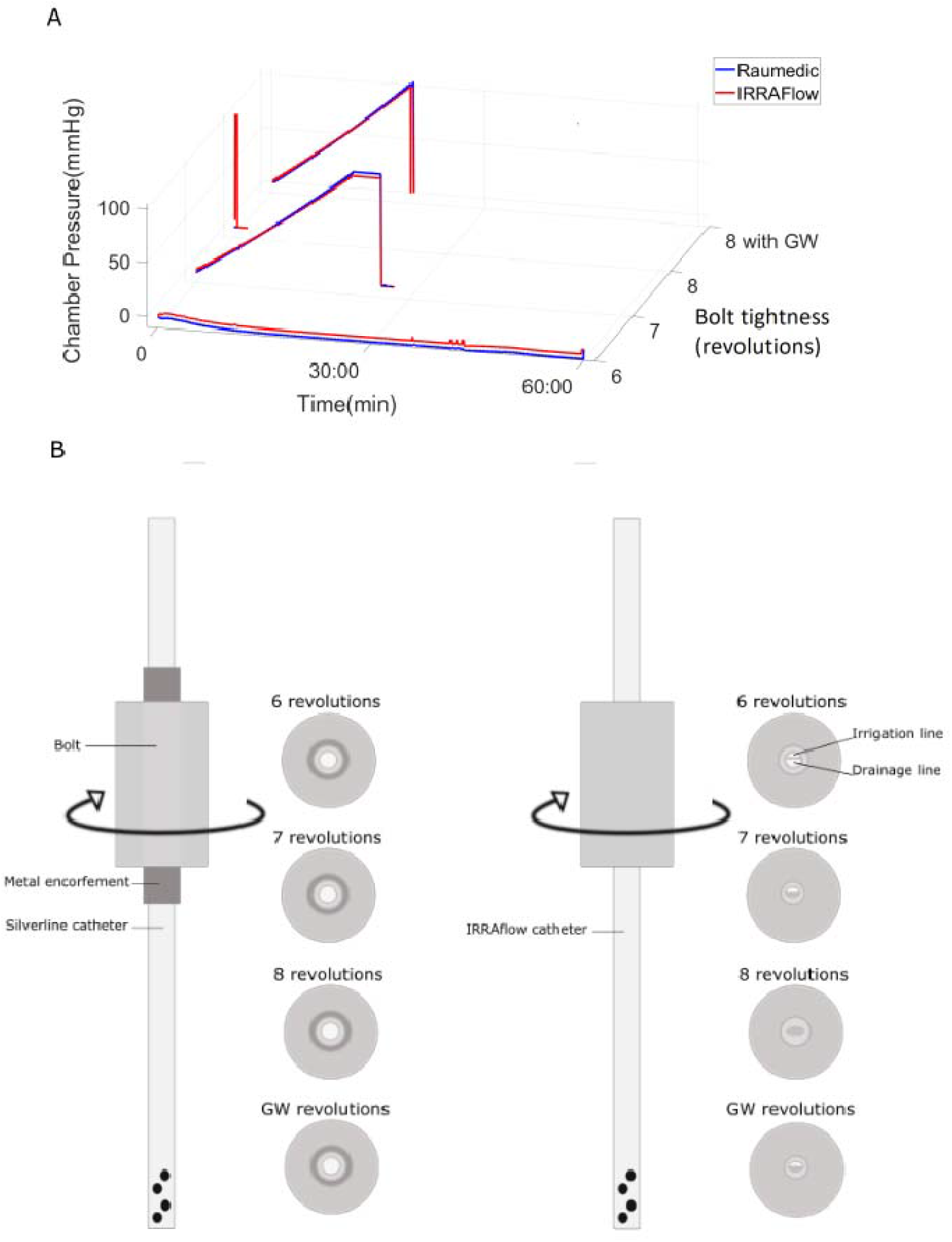
(A) Illustrates the pressure in the chamber under different bolt revolutions. ICP curves for 6, 7, 8 and stylet guided (Guide Wire, GW) revolutions on the bolt are shown. (B) Schematic figure showing the difference between the metal enforced Silverline® catheter (left) and the IRRA*flow*® dual lumen catheter (right). No metal reinforcement resulted in gradual compression and obstruction of the IRRA*flow*® catheter.

Our data showed that the IRRA*flow®* CPRS was obtained at varying sampling rates between 0.2 and 10 Hz. For example, during 1 second of irrigation, the sampling rate varied between 5 and 10 Hz. During 9 seconds of ICP measurement in the aspiration phase, the sampling rate was 0.2 Hz (Fig. 1). Further, the CPRS signal was characterized by a spike-type pressure increase during the irrigation phase followed by a rapid and stabilizing decay during the ICP measurement phase and a further drop during the aspiration phase (Fig. 1). The IRRA*flow®* pressure signal was obtained at a much lower sampling rate compared to the Raumedic device, which was sampled at 100 Hz.

### Experiment 1 – Bolt compression cap thightness test

This experiment was conducted to evaluate the performance of the catheter system at different bolt revolution settings. Results showed that at 6 revolutions or below, the catheter system did not exhibit constriction, and the integrity of the irrigation and drainage lines was preserved. However, we observed that the catheter was not properly fixed, retractable, and leaked during irrigation (Table 1, Fig. 2).

**Table 1.** Experiment 1 – Bolt compression cap tightness test Mean irrigation and drainage for the bolt constriction test.

| Bolt tightness | Irrigation | Drainage | Observations |
| --- | --- | --- | --- |
| 6 revolutions | 172 ml | 170 ml | Leakage observed |
| 7 revolutions | 64 ml | 0 ml | ICP increase, drainage compromised |
| 8 revolutions | 0 ml | 0 ml | Catheter fully constricted,<br>IRRAflow® ICP: 300 mmHg<br>Raumedic ICP: 0 mmHg |
| Maximal fastening with stylet in catheter (also 8 revolutions) | 58 ml | 0 ml | ICP increase, drainage compromised |

At 7 and 8 revolutions, we did not observe any leakage. However, at 7 revolutions, we observed constriction of the drainage line resulting in a progressive increase in chamber pressure, which reached 20 mmHg after 5 min and 30 sec and 99 mmHg after 28 minutes, indicating preserved irrigation performance but compromised drainage (Table 1, Fig. 2). At 8 revolutions, both irrigation and drainage lines were fully constricted. The chamber pressure was reliably 0 mmHg according to the Raumedic sensor. However, the IRRA*flow®* sensor simultaneously measured 300 mmHg, indicating significantly compromised and unreliable pressure sensing (Table 1, Fig. 2).

To alleviate the constriction, we attempted to secure the bolt around the catheter with the stylet (guidewire) in place and retracting the stylet before initiating irrigation. However, this option was not clinically feasible as the ICP reached 99 mmHg after 20 minutes. Therefore, the remainder of experiments was reliably performed at 6.5 revolutions, at which setting the chamber was tight, with no leakage, while irrigation/drainage was preserved, although the catheter was still easily retractable.

### Experiment 2 - IRRAflow® design space exploration

The results of the design space exploration are shown in Fig. 3. At a low irrigation (20 ml/hour), the optimal drainage bag level was 0 cm, resulting in nearly balanced fluid exchange with a mean net drainage of 5.7 ml (SE ± 1.77) and a mean chamber pressure of -1.78 mmHg (SE ± 0.23) over the course of 10 minutes. Conversely, over-drainage was observed at bag heights of -19 cm, -29 cm, and -39 cm. At 90 ml/hour irrigation positioning of the drainage bag at 0 cm or -19 cm resulted in mean net over drainage of 3.85 ml (SE ±1.7) and 3.04 ml (SE ±2.1), respectively, corresponding to end chamber pressures of -0.7 (SE ± 0.19) and 4.8 mmHg (0.23), respectively (Fig. 3).

**Figure 3.**
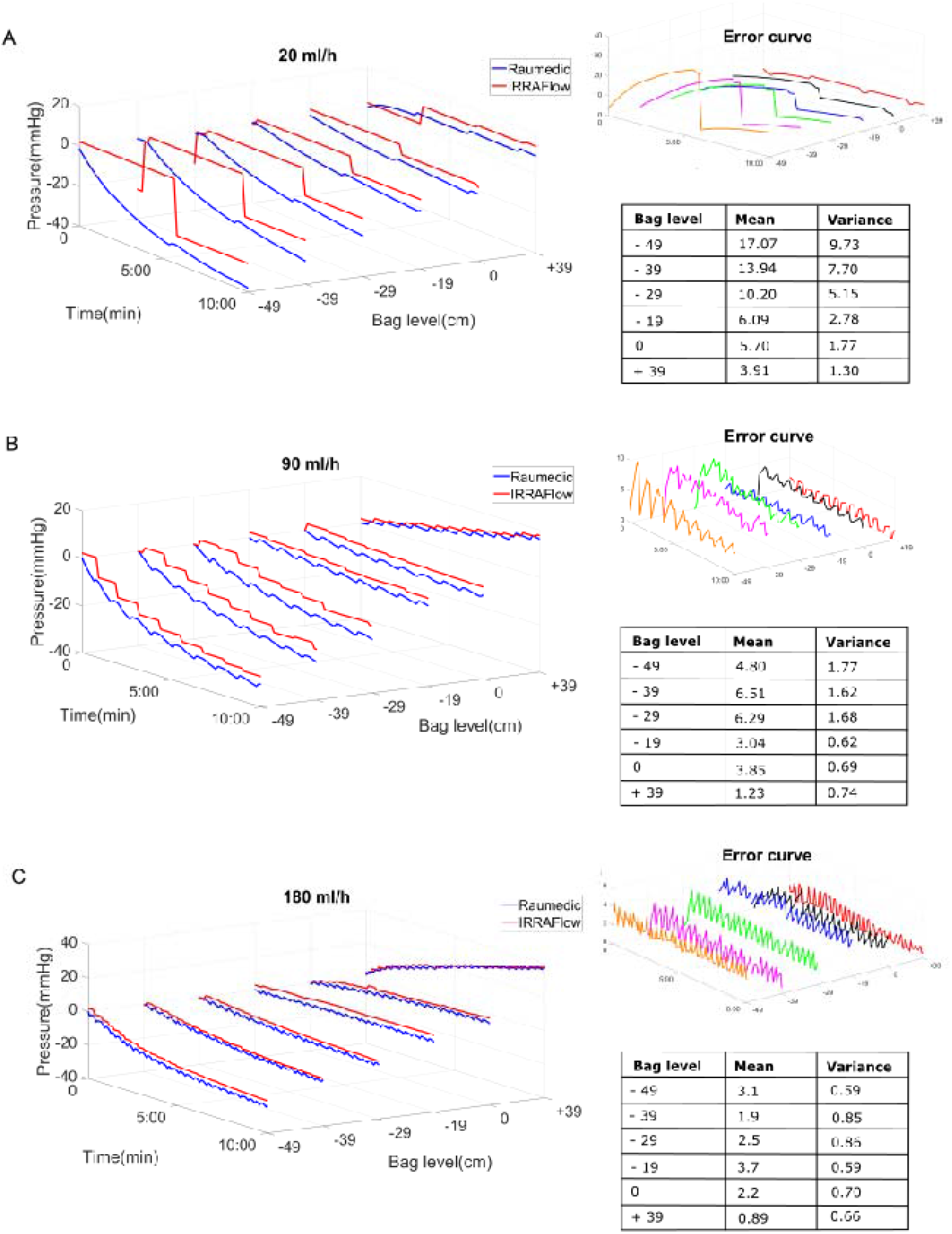
IRRA*flow®* settings and physiological implications. Pressure signals obtained at different IRRA*flow®* combinations of irrigation rate and drainage bag levels (A. 20/hour and 19 cm, 29cm and 39 cm, B. 90 ml/h and 19cm, 29cm and 39 cm and C. 180 ml/h and 19cm, 29cm and 39 cm (blue: Raumedic pressure signal, Red: IRRA*flow®* pressure signal). Error signals showing the pressure difference between the IRRA*flow®* and Raumedic pressure readouts are shown on the right side of each panel along with tables of the corresponding means and variances error in mmHg. The time required to achieve a stable chamber pressure varied with different settings and balanced fluid exchange required adjustment of drain bag to accommodate the irrigation rate.

At 180 ml/hour irrigation, the optimal drainage bag level was -29 cm, with a mean net over drainage of 2.5 ml (SE± 1.1) resulting in a slightly negative steady state end mean pressure of -6 mmHg (SE ± 0.39). At a drain bag height of -19 cm, slight underdrainage was observed (0.8 ml (SE ± 0.11) mean saline accumulation), resulting in a mean pressure increase of 6 mmHg (SE ± 1.98) (Fig. 3). The mean difference and variance between the ICP signals measured by IRRA*flow®* and the Raumedic devices is shown in Fig. 3 and in Supplementary Material 4. Overall, we found that the mean difference between the two signals varied between 0.89-17.07 mmHg (mean variance error 0.59-9.73 mmHg). This observation was mainly a result of under-sampling of the IRRA*flow®* ICP, which represented a single sample ICP obtained at the end of the ICP measurement phase which was then maintained throughout the drainage phase. The device therefore failed to capture and report the subsequent negative pressure that arose during the drainage phase.

### Experiment 3 - Accuracy of the IRRAflow® injection volume

The results from the IRRA*flow®* injection volume tests are shown in Figs. 4a and b. The observed ratio of change in baseline pressure is shown in Fig. 4. Bolus injections performed by the IRRA*flow®* device were stable and reliable in terms of volume at various chamber pressures. This reliability was validated by a stable pressure increase for each bolus and further a stable average mass of saline injected with each bolus (1.0061g/bolus, SE ± 0.059) compared with the anticipated mass of 1 g/bolus (equivalent to 1 ml/bolus as reported by the device 1000) (Supplementary material 4).

**Figure 4.**
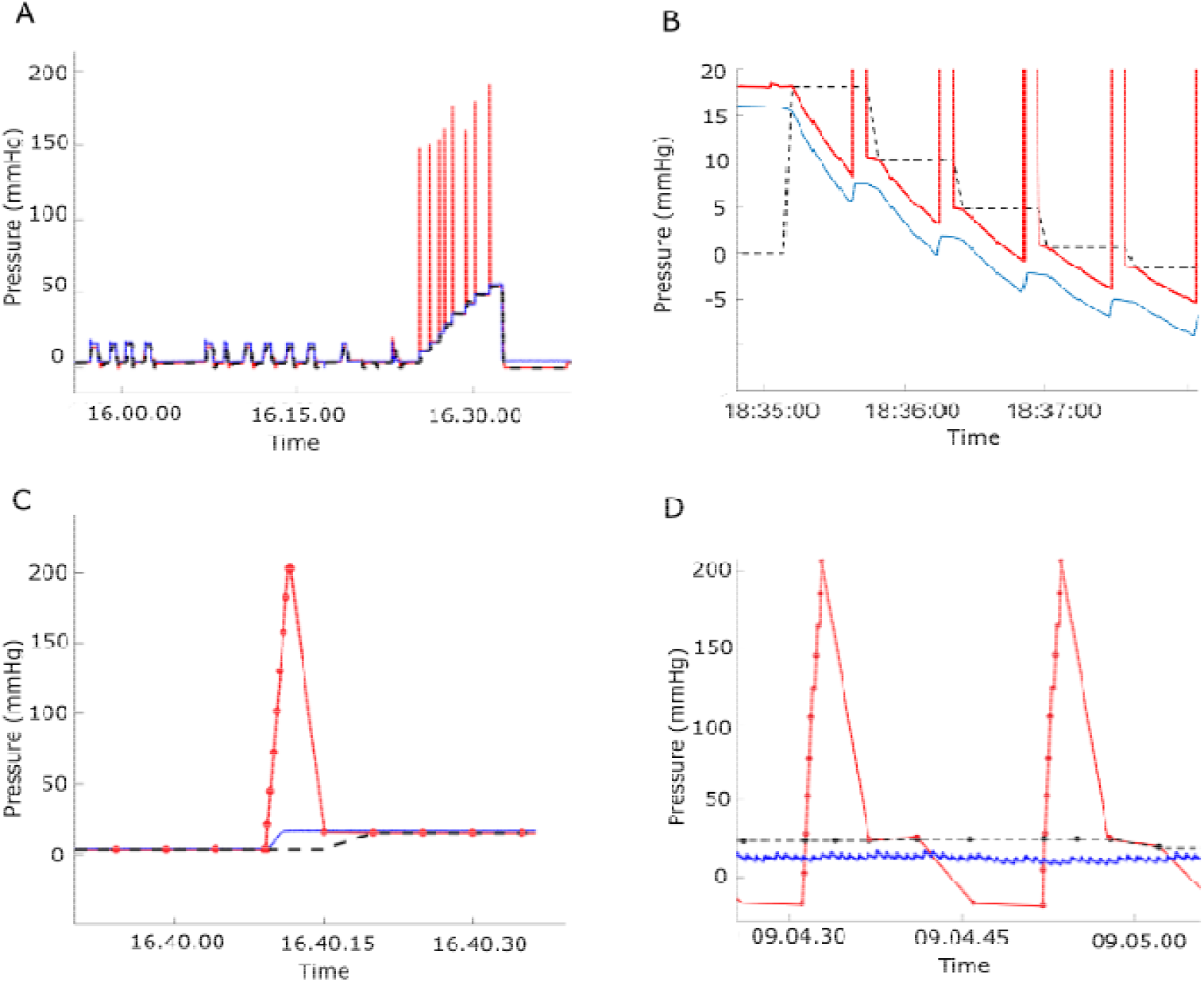
Reliability of the IRRA*flow®* pressure readout. Panel A shows spikes observed in the current pressure signal (CPRS) during the irrigation phase when the saline injections are performed by IRRA*flow®* as bolus injections (red). In comparison no spikes were observed following manual bolus injections using a syringe (blue). Panel B shows the negative pressure values during the aspiration phase. The blue line represents the Raumedic obtained signal and the red line represents the IRRA*flow®* obtained signal. Panel C shows a representation of the IRRA*flow®* (CPRS and ICP) and Raumedic pressure signals, respectively, obtained from the ex vivo model Infusion results in a clear pressure increase of 200 mmHg, which is consistent across all signals. In comparison, panel D shows similar data obtained from a patient. Contrary to the ex vivo data in panel C, we observed no increase in pressure in the ICP upon bolus infusion. This was thought to be partly due to the low signal to noise ratio (SNR), given the ICP variations due to the heart beat, but also likely related to a lack of pressure transmission from the catheter tip to the Raumedic device. Panel D shows that no negative pressures are observed in the Raumedic data (blue) during the aspiration phase as is observed in the IRRA*flow®* data (red line during aspiration phase).

### Experiment 4 - Accuracy and reliability of the IRRAflow® pressure sensor

During the irrigation phase, we observed spikes in the CPRS signal *ex vivo* with high amplitudes of up to 200 mmHg (Fig. 4). These values were approximately 15 times higher than the corresponding ICP value measured using the Raumedic device (Fig. 4). The CPRS then decayed exponentially throughout the subsequent 9 seconds ICP measurement phase to reach a nearly stable plateau at the end. However, during the decay phase, the signal was only sampled twice. Contrary to the IRRA*flow®* CPRS, the Raumedic ICP signal did not exhibit spike features following saline injection with IRRA*flow®*. Rather, we observed an expected pressure increase (8 mmHg, SE ± 4.2) for all three pressure signals reaching a stable plateau, which was maintained throughout the rest of the ICP measurement phase (Fig. 4C). We observed no spikes in the CPRS or IRRA*flow®* signals during manual 1 ml bolus injection using a syringe indicating that the spikes were an irrigation artifact inherent to the IRRA*flow®* system (Fig. 4).

During the aspiration phase of the IRRA*flow®* cycle, we observed a decay of the CPRS and Raumedic signals *ex vivo* to reach negative values between -7 and -13 mmHg (Fig. 4), reflecting the active aspiration of liquid out of the chamber during this phase (Fig. 4B). A similar pressure decay in the CPRS signal was observed in the patient data, although it was not transmitted to the Raumedic ICP sensor (Fig. 4D). This was thought to be partly due to the low signal to noise ratio (SNR), given the ICP variations secondary to the heartbeat, but also likely related to a lack of pressure transmission from the catheter tip to the Raumedic device.

In all experiments, we observed discrepancies between the IRRA*flow®* CPRS signal and the corresponding Raumedic ICP signal. In the idealized *ex vivo* setting, the mean deviation was 3.8 mmHg (SE ±2.6) (Fig. 5), while the synchronized data set retrieved from two real patients treated with IRRA*flow®* had periodical deviations with IRRA*flow®* ICP values up to 10-15 mmHg higher than values measured using Raumedic (mean 12.0 mmHg, SE ± 3.6 mmHg).

**Figure 5.**
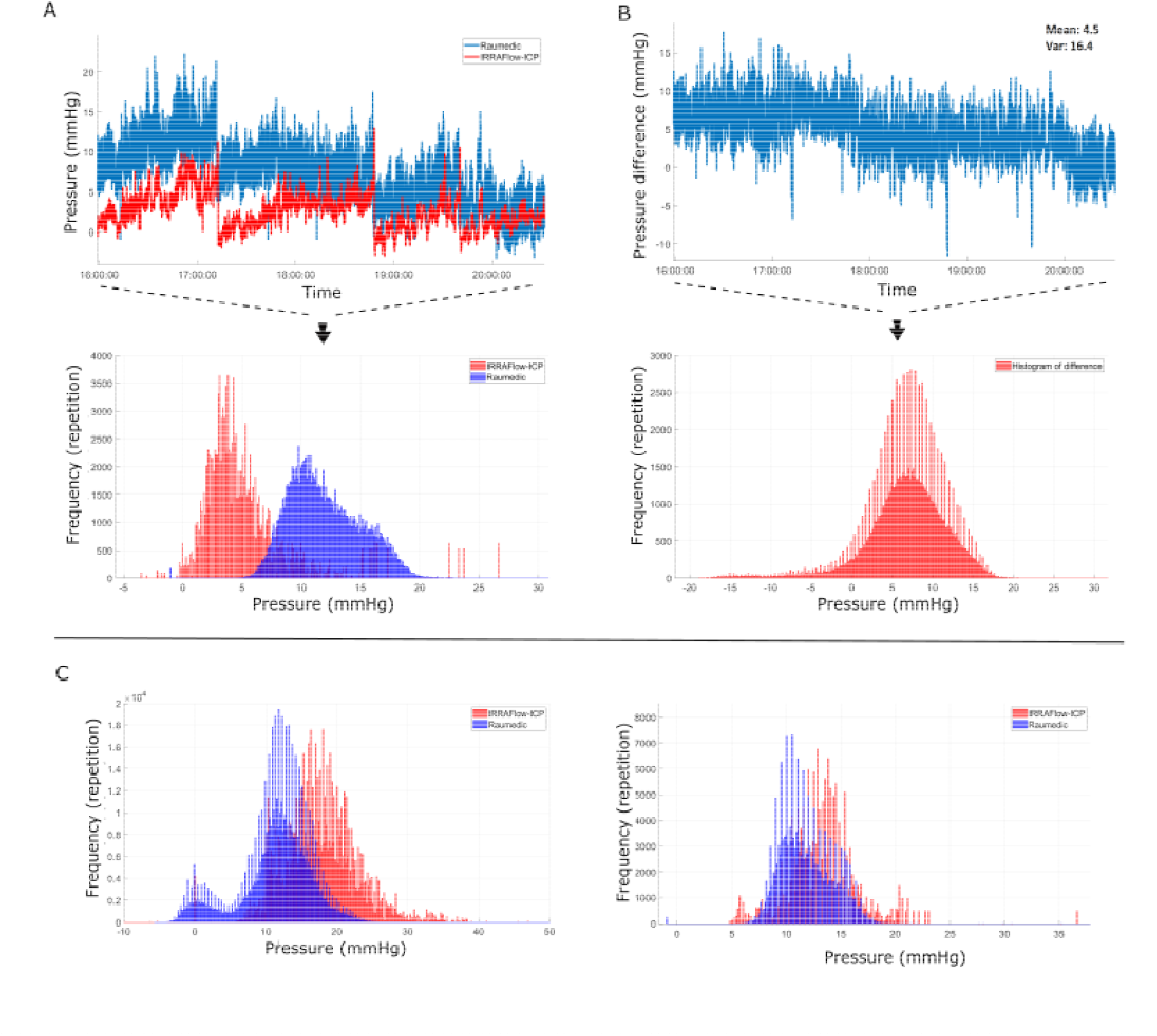
Discrepancies in the IRRA*flow®* vs Raumedic obtained pressure signals. A. Synchronized patient data. The upper curve shows the signal recorded during 6 hours from patient 3 (red: IRRA*flow®* and blue: Raumedic) and below the corresponding histogram of the synchronized ICP signals from patient 3 and 4 . B shows the difference between the two signals (mean difference 4.5 mmHg and variance 16.4 mmHg) and the corresponding histogram of the difference in ICP signals (mean 3.8 mmHg, SE 2.6, p<0.001). C. shows histograms of unsynchronised ICP signals from patient 1 and 2 (mean difference 6.31 mmHg, variance 28.6, p<0.0001).

In addition, the unsynchronized dataset revealed a significant difference in the ICP values comparing the IRRA*flow®* ICP values to the Raumedic ICP values (patient 1: mean difference 6.31 mmHg, variance 28.6, p<0.0001 and patient 2: mean difference 1.41 mmHg, variance 8.29, p<0.0001, normally distributed) (Fig. 5) with the IRRA*flow®* values higher than Raumedic values. Importantly, the error was periodically very high, with significant deviations up to 19 mmHg between the two devices. These periodical errors in ICP measurement were often related to catheter occlusions if the catheter tip was placed in blood or had displaced into the brain parenchyma and not surrounded by liquid.

Furthermore, we observed a significant mean drift difference of 3.16 mmHg (variance 0.4 mmHg, p=0.05) over a 24-hour test period with a mean 24-hour drift of 3.66 mmHg (Var 0.28 mmHg) in the IRRA*flow®* measured pressures compared to 0.5 mmHg (variance 1.12 mmHg) in the Raumedic measured pressures (Fig. 6).

**Figure 6.**
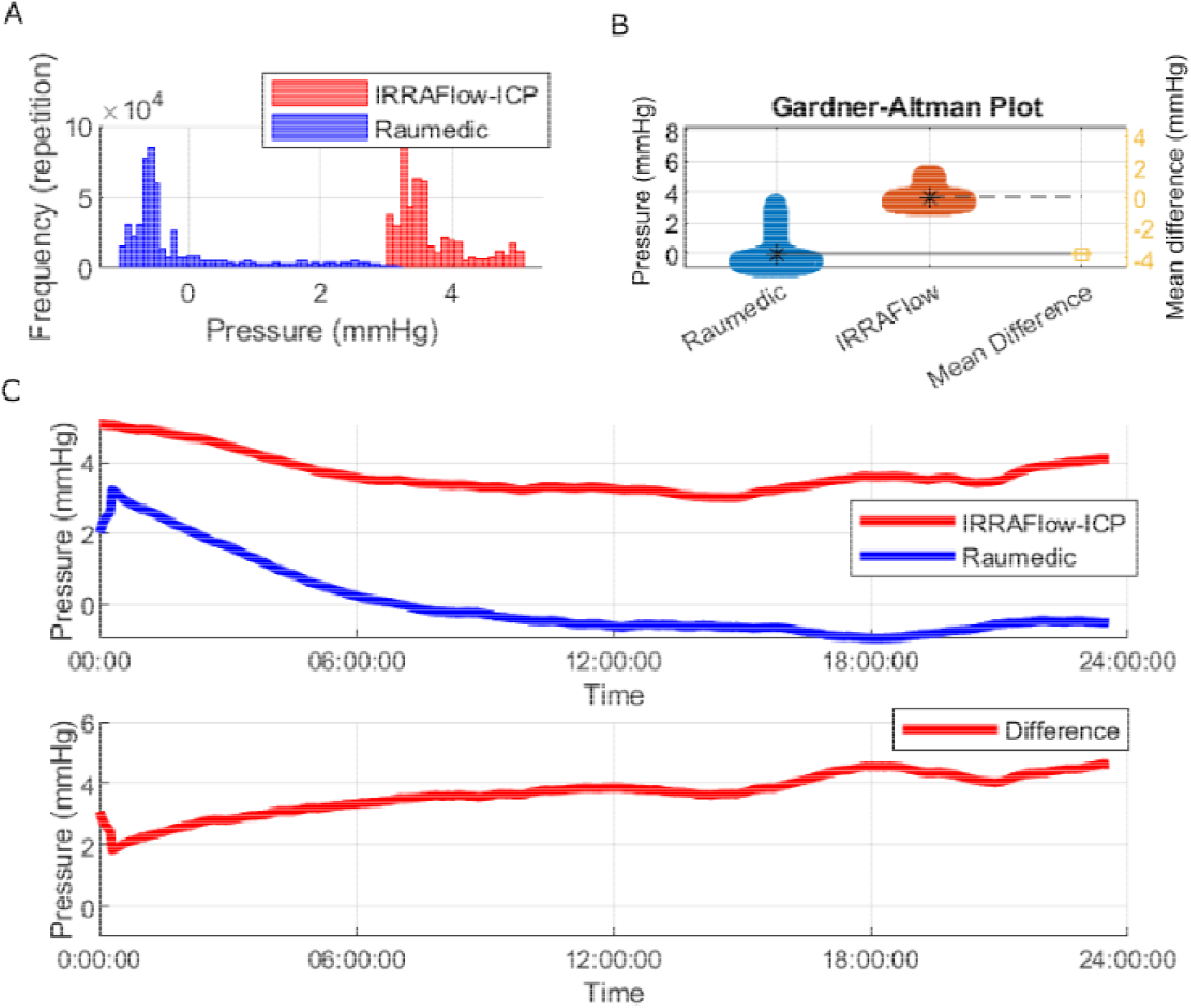
24 hour drift tests. Panel A shows a histogram of the recorded drift data from the Raumedic (blue) and the IRRA*flow®* (red). Panel B shows the Gardner-Altman plot and the mean difference of the recorded signals. Panel C shows the 24 hours drift test recording and below the difference is shown.

Lastly, we observed a low signal to noise ratio (SNR). During 1 second of injection in the irrigation phase the frequency of the signal was found to be overlapping with the heartbeat in the in vivo data. In the *ex vivo* model, with the absence of heartbeat, changes in pressure during 1 second of irrigation with 1 ml of saline were easily observed (Fig 4C). The same change in pressure was not observed in the patient data (Fig. 4B).

## Discussion

IRRA*flow®* is a new and potentially promising technology for automated and controlled irrigation/aspiration treatment of IVH using tunneled dual-lumen silicone catheters. Currently, there are no clinical guidelines describing how to implement and set up the IRRA*flow®* device for approved indications, including IVH. In this study, we conducted a comprehensive assessment of the implications and effects of different IRRA*flow®* settings as well as an evaluation of the reliability of IRRA*flow®* volume injections and pressure measurements. Our study was based on an idealized ex vivo setup as well as patient data.

Previous studies have established that bolt fixation of external ventricular catheters is associated with reduced risk of catheter displacement, CSF leakage and CNS infection. For that reason, it is potentially compelling to use conventional and approved bolt EVD technologies to fixate the IRRA*flow®* catheter during treatment. However, our study showed that this approach is indeed not safe or feasible, when combining the IRRA*flow®* catheter (version 2.0 ICGS020) with the widely used Spiegelberg Silverline*®* bolt, as this was associated with improper fixation, leakage, or catheter constriction. This combination represents a patient hazard and should be avoided.

We evaluated a variety of device settings to determine the optimal settings for clinical implementation. We found that irrigation was associated with a corresponding adjustment of the intracranial pressure until a steady state was reached with balanced infusion and aspiration. The steady state pressure was determined by the drain bag height, which could be adjusted to stabilize the ICP at the baseline value and accommodate different irrigation rates. As such, higher irrigation rates should be followed by lower drain bag heights to create a higher negative aspiration gradient during the shortened drainage period, to achieve a balanced saline influx and efflux. Specifically, we found that a near-balanced perfusion was obtained at the following settings:

- Irrigation rate 20 ml/h = drainage bag at 0 cm
- Irrigation rate 90 ml/h = drainage bag at 19 cm
- Irrigation rate 180 ml/h = drainage bag level at 29 cm

For clinical implementation the drain bag height could be adjusted to achieve a stable ICP in the desired range for any given irrigation rate. Elevated drainage bag heights may result in net intracranial CSF accumulation and corresponding undesired ICP elevation. Similarly, very low drainage bag levels may result in severe over drainage. Low drainage bag levels potentially result in very negative pressure gradients during the aspiration phase. This may be associated with an increased risk of undesirable aspiration of ependyma, or blood clots followed by catheter occlusions, and we recommend to avoid very negative drainage bag levels. The negative pressure gradient during aspiration is not displayed to the clinician on the IRRA*flow®* device, which only reports the point sampled ICP before the drainage valve opens. This represents an important pitfall and a potential for improvement of future devices. To evaluate the reliability of the internal IRRA*flow® ICP monitor,* we compared the pressure readouts to those of a Raumedic Neurovent P ICP sensor. We found that the IRRA*flow®* pressure sensor, which is located proximally to the catheter tip, is prone to significant spike artifacts resulting from saline infusion and subsequent pressure stabilization. The pressure significantly declines during the subsequent aspiration phase, and the estimated ICP does not continuously represent the true ICP throughout the cycle. The sensor requires steady state conditions with no liquid movements in the catheter to be accurate. Furthermore, it requires unobstructed passage of liquid, which potentially compromises accuracy during patient treatment, e.g., due to clot obstruction or collapse of the ventricular system during aspiration with negative gradients, as described above. This was indeed reflected in significant deviations between the IRRA*flow®* pressure data and the Raumedic pressure data obtained from four IVH patients. Particularly, we observed significant periodical deviations with faulty ICP readouts using IRRA*flow®*, which we believe are associated with the technical design as described above. This represents a significant problem as the IRRA*flow®* operational cycle is governed and supervised by this internal pressure monitor. False pressure readouts may therefore result in unsafe device operation or frequent alarms. For this reason, we recommend that an additional parenchymal ICP sensor be used in conjunction with IRRA*flow®* to ensure stable and reliable pressure sensing during irrigation. In the idealized ex vivo setup, we observed only minor deviations between the pressure readouts from the IRRA*flow®* and Raumedic sensors. The extent of sensor drift was on average higher for IRRA*flow®*, which may indicate a need for regular and frequent calibration. The American National Standard Institute (ANSI)/the Association for the Advancement of Medical Instruments (AAMI) has recommended that ICP monitors should be reliable within the ICP range of 0-100 mmHg. Especially in the ICP range of 0-20 mmHg, the drift should be within ± 2 mmHg(6). However, in the experiment we compared a catheter tippet ICP monitor to an intraparenchymal probe which could be a limitation to the drift test. For future systems, we recommend that the pressure sensor be integrated into the catheter tip to avoid artifacts and more accurately represent ICP continuously.

We observed negative pressure values in the IRRA*flow®* and Raumedic data in the *ex vivo* model but only in the IRRA*flow®* data retrieved from the patient (Fig. 4). This finding could be due to a local decrease in pressure around the catheter tip only, with no affection of the pressure in the parenchyma where the Raumedic probe is located.

We believe that the problems observed with proximal pressure sensing in the IRRA*flow®* control unit are related to the analogous behavior of IRRA*flow®* to a resistor, inductance, and compliance (RLC) circuit (Fig. 1). The cycle of the IRRA*flow®* is designed to start with a phase of irrigation followed by a phase of ICP measurement and again followed by a phase of aspiration (Fig. 1). During ICP measurement the fluid in the tubes has no movement and the same pressure was observed in the IRRA*flow®* control unit and the Raumedic device. However, when irrigation and aspiration were initiated, the pressure differed in the two devices (Fig. 1, PI and PR).

## Conclusion

Near-balanced irrigation can be achieved by adjusting the IRRA*flow®* drainage bag to accommodate different irrigation rates. It is not possible to provide a clear guideline to define the appropriate settings. internal IRRAflow*®* ICP monitor is based on an unfavorable proximal pressure-sensor technology and is unreliable compared to the Raumedic. We recommend that use of the IRRA*flow®* device is accompanied by a separate ICP monitor for improved patient safety. The IRRA*flow®* volume injections are reliable at all physiological ICP values.

## Data Availability

All data produced in the present work are contained in the manuscript

