## Supplementary 1-4 for "Reliability and Performance of the IRRAflow® System for Intracranial Lavage and Evacuation of Hematomas - A Technical Note": Supplementary material lab test .docx

**Supplementary material 1**. a and b showing the reverse tunneling and fastening of the IRRA*flow*® catheter. Figure c showing the bolted standard passive external ventricular drain (EVD), Speigelberg, Silverline®.

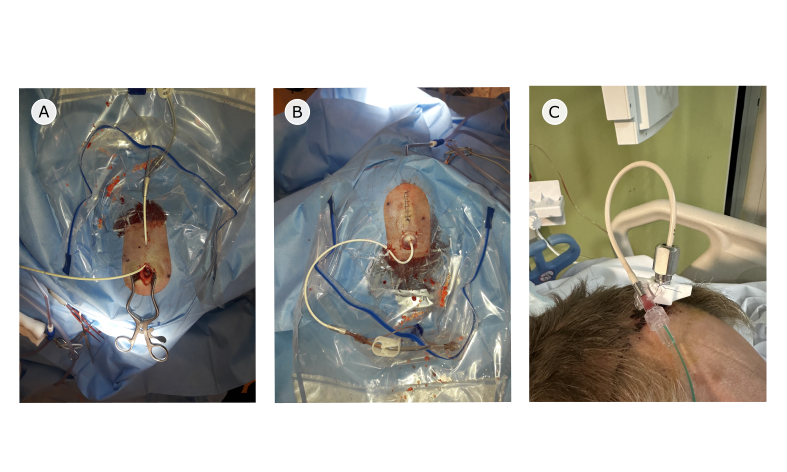

**Supplementary material 2.** Lab set up in Compliance test, experiment 1 and 2

| **Camber compliance test** |  |
| --- | --- |
| Room temperature (°celcius) | 23°celcius |
| Water in artificial brain (ml) | 100-600 ml (variable) |
| IRRAflow® lower alarm (mmhg) | Device not in use |
| IRRAflow® upper alarm (mmhg) | Device not in use |
| IRRAflow® drain above (mmhg) | Device not in use |
| IRRAflow® irrigation level (ml/hour) | Device not in use |
| IRRAflow® drainage bag level (cm) | Device not in use |
| Experiment repetitions | 5 times |
| **Experiment 1 – Bolt constriction test** |  |
| Room temperature (°celcius) | 23°celcius |
| Water in artificial brain (ml) | 200 ml |
| IRRAflow® lower alarm (mmhg) | - 99 mmhg |
| IRRAflow® upper alarm (mmhg) | + 99 mmhg |
| IRRAflow® drain above (mmhg) | - 99 mmhg |
| IRRAflow® irrigation level (ml/hour) | 180 ml/h |
| IRRAflow® drainage bag level (cm) | 29 cm |
| Duration of each experiment | 1 hour |
| Experiment repetitions | 5 times |
| **Experiment 2 – IRRAflow design space exploration** |  |
| Room temperature (°celcius) | 23°celcius |
| Water in artificial brain (ml) | 200 ml |
| IRRAflow® lower alarm (mmhg) | - 99 mmHg |
| IRRAflow® upper alarm (mmhg) | + 99 mmHg |
| IRRAflow® drain above (mmhg) | - 99 mmHg |
| IRRAflow® irrigation level (ml/hour) | Variable (20 ml/h, 90 ml/h and 180 ml/h) |
| IRRAflow® drainage bag level (cm) | Variable (+39 cm, 0 cm, -19, cm, -29 cm, -39 cm -49) |
| Duration of experiment at each setting | 10 minutes |
| Experiment repetitions | 5 times at each setting |
| **Experiment 3 - Accuracy of the IRRAflow injection volume** |  |
| Room temperature (°celcius) | 23°celcius |
| Water in artificial brain (ml) | 200 ml |
| IRRAflow® lower alarm (mmhg) | - 99 mmHg |
| IRRAflow® upper alarm (mmhg) | + 99 mmHg |
| IRRAflow® drain above (mmhg) | - 99 mmHg |
| IRRAflow® bolus setting | 1 ml |
| **Experiment 4 - Accuracy and reliability of the IRRAflow pressure sensor** |  |
| Room temperature (°celcius) | 23°celcius |
| Water in artificial brain (ml) | 200 ml |
| IRRAflow® lower alarm (mmhg) | - 99 mmHg |
| IRRAflow® upper alarm (mmhg) | + 99 mmHg |
| IRRAflow® drain above (mmhg) | - 99 mmHg |
| IRRAflow® bolus setting | 1 ml |

**Supplementary material 3.** Data output from the IRRAflow control unit. (SysS: System status , TrtS: treatment status, ICP: intracranial pressure, Cprs: current pressure, HLim: Upper alarm limit, LLim: Lower alarm limit, PM: Preset Mode (1 or 0), (system operating on default settings or not, PPrs: Preset pressure (drain above), Ivol: infusion volume (ml/hour), CycT: cycle time (sec), stat1 HEX printout of various system status, Stat2: HEX printout of various system status, ErrCode: Errorcode)

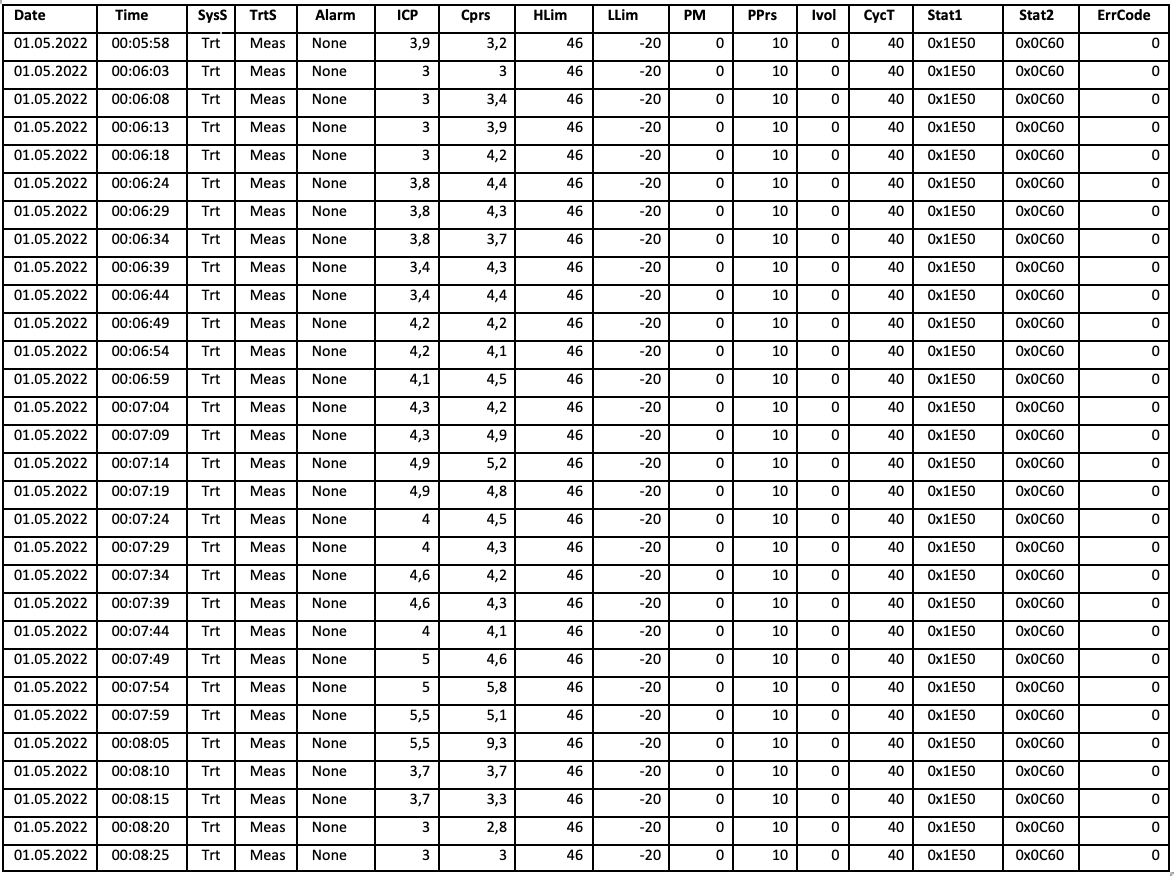

**Supplementary Material 4.** A. Change in baseline pressure following IRRAflow® bolus injections and B. table showing the mean actual injected volume at each bolus injection in grams.

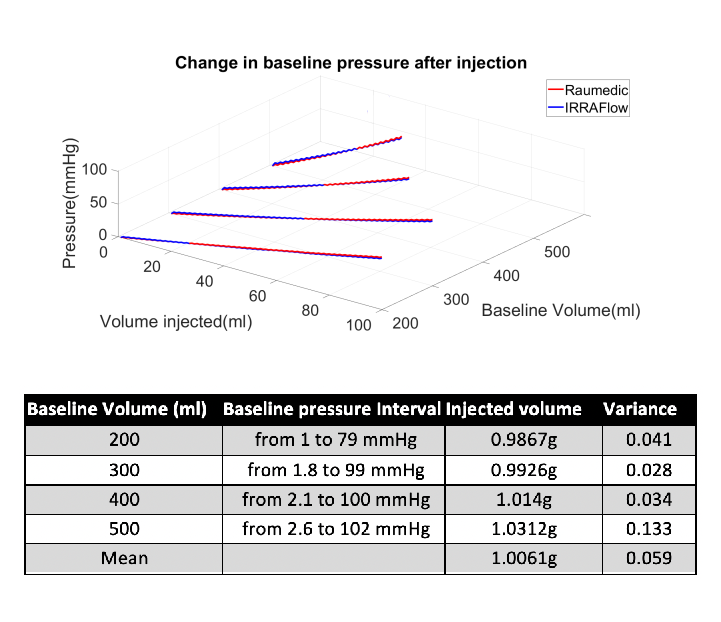
